# Precision association of lymphatic disease spread with radiation-associated toxicity in oropharyngeal squamous carcinomas

**DOI:** 10.1101/2020.08.25.20181867

**Authors:** Andrew Wentzel, Timothy Luciani, Lisanne V. van Dijk, Nicolette Taku, Baher Elgohari, Abdallah S. R. Mohamed, Guadalupe Canahuate, Clifton D. Fuller, David M. Vock, G. Elisabeta Marai

## Abstract

**Purpose:** Using a cohort of 582 head and neck cancer patients with nodal disease, we employed clustering over a novel graph-based geometrical representation of lymph node spread in order to identify groups of similar patients. We show that these groups are significantly correlated with radiation-associated dysphagia (RAD), and predictive of late aspiration and feeding tube toxicity.

**Materials and methods:** All patients received radiotherapy for oropharyngeal cancer (OPC) and had non-metastatic affected lymph nodes in the head and neck. Affected lymph nodes were segmented from pretreatment contrast-enhanced tomography scans and categorized according to consensus guidelines. Similar patients were clustered into 4 groups according to a graph-based representation of affected lymph nodes. Correlation between dysphagia associated symptoms and patient groups was calculated.

**Results:** Out of 582 patients, 26% (152) experienced toxicity during a follow up evaluation 6 months after completion of radiotherapy treatment. Patient groups identified by our approach were significantly correlated with dysphagia, feeding tube, and aspiration toxicity (p <.0005). Conclusion: Our work successfully stratified a patient cohort into similar groups using a structural geometry, graph-encoding of affected lymph nodes in OPC patients, that were predictive of late radiation-associated dysphagia. Our results suggest that structural geometry-aware characterization of affected lymph nodes can be used to better predict radiation-associated dysphagia at time of diagnosis, and better inform treatment guidelines.

## Introduction

Despite an overall reduction in tobacco and alcohol associated malignancies of the head and neck cancers (HNC), recent decades have been marked by a paradoxical increase in the incidence of cancers of the oropharynx (OPC) [1]. Approximately 7,500 cases of OPC are diagnosed in the United States annually, including 70-90% of which are associated with the human papilloma virus (HPV) [2-4]. HPV-related OPC patients are more likely to be young (i.e. < 55 years old), and generally have better prognosis than HPV-negative OPC patients, with 5- year overall survival rates around 80%, compared to rates of less than 50% for HPV-negative patients [5]. The steady increase of OPC survivors has amplified the need to understand and minimize acute and long term side effects following (chemo-)radiation based on cohorts of similar patients [6]. Swallowing dysfunction, or radiation-associated dysphagia (RAD), has a particularly large impact on the quality of life of OPC survivors [7]. RAD can result in nutritional deficiencies and life-threatening aspiration pneumonia [8]. Over 50% of patients with locally-advanced OPC will demonstrate acute or subacute evidence of aspiration and nearly 10% will become gastrostomy tube dependent during the peri-treatment course [9-12].

Studies have shown that the maximum distance between involved lymph nodes and primary tumors are potential determinants of metastasis-free survival, and that patterns of involved (diseased) lymph nodes (LN), as well as tumor proximity to organs at risk, may affect toxicity [13,14]. However, matching spread patterns based on lymph labels alone and ignoring spread over adjacent anatomical regions has been shown to result in incorrect patient matches [15]. These data suggest that spatial information pertaining to lymph node involvement may be useful in predicting patient outcomes, and inclusion of stratifications derived from complex data into risk prediction models for OPC patients has proven effective in improving model performance [16]. Given the overall favorable survival outcomes of HPV-associated disease, there is a need for staging tools that can integrate baseline clinical information and stratify patients by risk for treatment-related, clinical toxicities. We hypothesize that the pre-treatment pattern of involved lymph nodes can be used to predict RAD in OPC patients. To evaluate this hypothesis, we propose a novel patient risk stratification based on the spatial similarity of the affected lymph node patterns. Concretely, we encode chains of affected lymph nodes as a set of covariates that incorporate spatial relationships between affected nodes and apply unsupervised clustering to stratify patients into 4 groups. The resulting groups are strongly correlated with 2 post-treatment toxicities associated with RAD, and can serve as a granular toxicity risk-staging system based on nodal disease spread.

## Material and Methods

An IRB-approved, retrospective review of patients diagnosed with oropharyngeal cancer (OPC) and treated at MD Anderson Cancer Center from 2005 to 2013 was performed. Patients with lymph node positive, biopsy-proven, OPC who were treated with radiotherapy (RT) with or without chemotherapy with curative intent and had at least 6 months post-treatment follow-up assessment were eligible for inclusion.

Patient demographics, clinical data, outcomes, and toxicity assessments were collected retrospectively from electronic medical records. All patients underwent a complete physical and endoscopic examination, as well as radiological and pathological assessments at initial treatments. Data on affected lymph nodes levels were taken from baseline contrast-enhanced tomography (CECT) scans obtained during initial assessment. Lymph node levels extracted from CECT scans were used to construct a map of each patient’s lymphatic disease spread. LN levels were defined based on anatomical landmarks. LN levels considered were the retropharyngeal larynx (RPN) and levels I-VI based on consensus guidelines [17]. Outcome data was collected during a 6-month follow-up assessment that included physical, endoscopic, radiolocal, nutritional and toxicity assessments. Radiation associated dysphagia was defined as the presence of grade 2+ aspiration rate based on CTCAE guidelines [18], or feeding-tube insertion during treatment or after treatment completion. No feeding tubes were placed prophylactically.

To perform stratification, each patient’s chain of affected lymph nodes was encoded as a multidimensional vector (Appendix A). First, a map of levels I-VI, and the RPN, were constructed such that anatomically adjacent regions were connected in the map (Figure 1). Involved lymph nodes in each patient were identified for both sides of the head through physical examination and radiological imaging [19,20]. If at least one lymph node in a given level is affected with cancer cells, radiation oncologists refer to the corresponding node level as being involved with disease, and they involve the whole node level in treatment. Involvement was treated as separate covariates for the side of the head with the primary tumor (ipsilateral) and the side opposite the primary tumor (contralateral). When the primary tumor (GTVp) crossed the midline of the head (bilateral), the side with the larger bulk of primary disease was treated as the ipsilateral side. Figure 1 illustrates the encoding process for an example patient with bilaterally affected levels IIA-IIB and unilaterally affected level 3.

**Figure 1.**
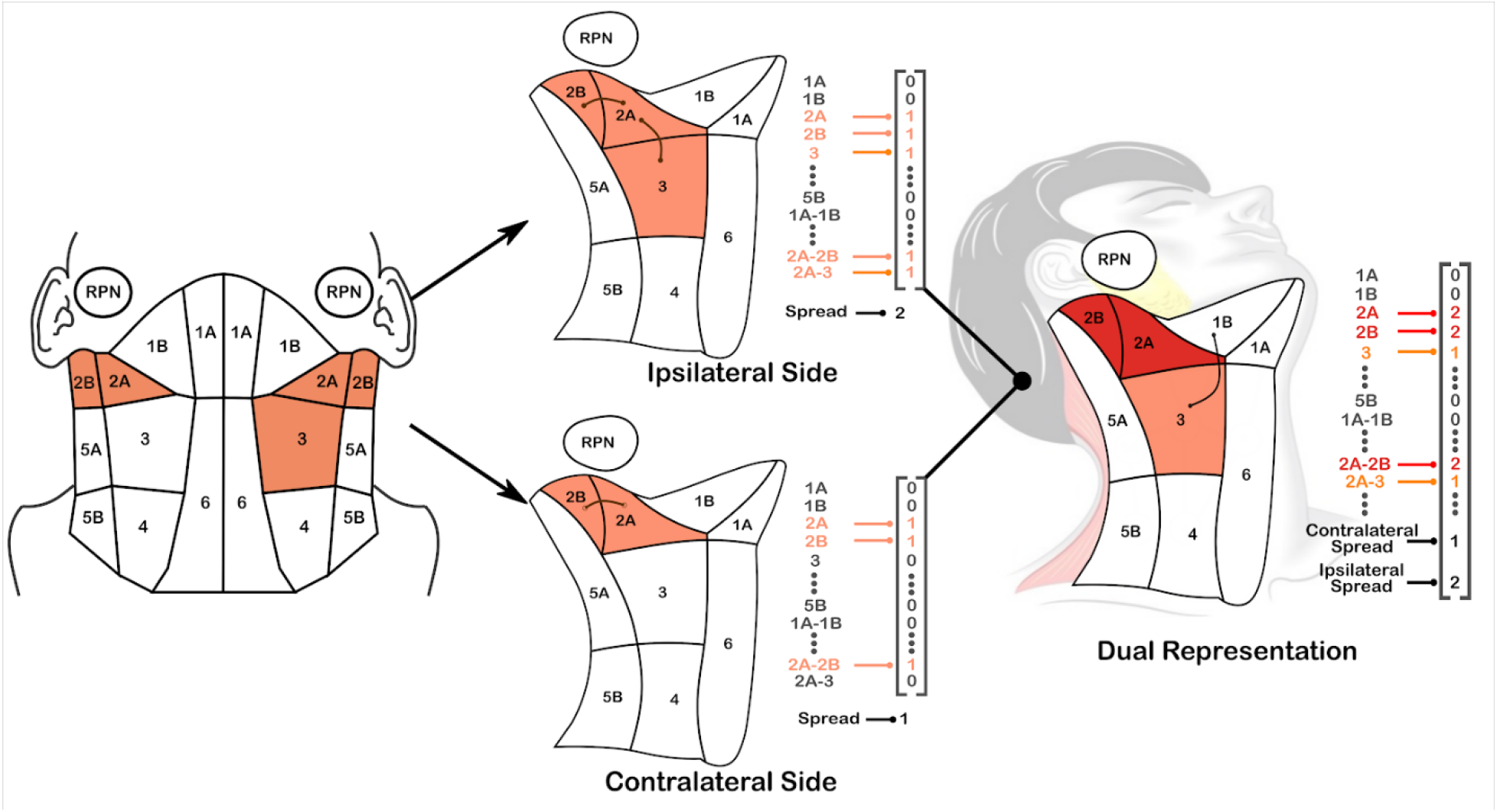
(Left) Diagram of the anatomically-adjacent representation of affected lymph nodes, for an example patient with levels 2A-2B affected on both sides of the head and level 3 affected on the ipsilateral side of the head. Adjacent regions are represented as bigrams (e.g., 2A-2B) in our encoding (Appendix A). (Center) Encodings for each side of the head. The contralateral side is given a ‘maximum spread’ value of 2 to represent the travel distance from 2b to 3, while the contralateral side (opposite primary tumor) has a spread of 1 for 2A-2B. (Right) Composite encoding for the patient that captures symmetry of the head and neck; diseased regions are added while the maximum LN spread on each side are treated as two separate covariates.

Pairwise similarity between patients was calculated using similarity based on LN spread over adjacent anatomical regions, which is an extension of the methodology introduced by Luciani et al. [15]. Their work showed that including anatomical adjacency information provides clinically-valid groupings of similar patients, whereas ignoring anatomical information does not. Stepwise hierarchical agglomerative clustering (HAC) was used to group similar subjects in the cohort as follows: First, the encodings corresponding to each unique spread pattern in the cohort were calculated. Values were then normalized across the identified patterns to have a mean of 0 and standard deviation of 1. Clustering was performed using Ward’s method [21] with the squared Canberra distance metric [22]. Ward’s linkage method minimizes the within-cluster sum of squared deviations from points to centroids, yielding more balanced clusters than other linkage methods. Canberra distance is a weighted version of Manhattan distance that allows us to account for the heterogeneity in the number of lymph node involvement between patients. We then mapped the pattern clusters back to the original cohort to identify cluster membership for the original patients.

In order to determine the optimal number of clusters to use, cluster labels were generated for 2 to 5 clusters. Predictive models of toxicity 6-months after finishing radiation treatment were created using different numbers of clusters, and compared to models based on N-staging, to allow for comparison against existing nodal risk categories. The optimal number of clusters was chosen using 10-fold cross validation and a logistic regression model for toxicity, further described below. Statistical analysis and a descriptive characterization was performed on the groups to show that the resulting clusters are correlated with RAD, and provide measurable improvement to predictive models using existing diagnostic information. In our reports below, clusters are labeled in the order of percentage of patients with RAD, where cluster 4 had the highest risk.

The cluster labels were used to create a logistic regression model to predict either feeding tube or aspiration (i.e. RAD), and evaluated using the area-under-curve (AUC), also known as the c-statistic, which is a measure of the effectiveness of the rankings, based on how the specificity of the prediction changes as the sensitivity is adjusted.

Fisher’s exact test [23] was used to measure the correlation between the resulting clusters and either post-therapy aspiration rate or feeding tube insertion. As Fisher’s exact test is not feasible on such large covariate sets, the Pearson Chi-square test was used to measure the clusters’ correlation with a set of baseline clinical covariates: T-category (primary tumor staging) and N-category (nodal tumor staging), HPV (p16+) status, total GTVp dose of at least 70 Gy, tumor subsite, primary tumor laterality, 7th edition AJCC staging, and age bracket (<= 55 years, 55-65 years, and > 65 years old).

Furthermore, to assess whether or not the LN clusters are significantly associated with RAD after accounting for known covariates, logistic regression models were trained to predict RAD using age (<65 vs 65 years or older), smoking status (never vs. current and former), use of chemotherapy (concurrent or combination), HPV status (positive, negative or unknown), T-category (1 or 2, vs 3 or 4), and N-category (1 or 2, vs 3 or 4). This baseline model was then compared to one with our LN clusters added in, and both models were tested using a likelihood ratio test to assess if the performance of the model with our LN clusters are significantly different (and better) than the baseline.

LN graph calculations and patient similarity computations were performed in Python using the pandas library [24]. Hierarchical clustering and statistical tests were performed using Matlab 2018a statistical toolbox and R. All statistical tests were two-sided with p < 0.01 considered statistically significant.

## Results

Out of 644 OPC patients available for the study, 582 patients had affected lymph nodes and were included in the final cohort. Patients’ demographics, clinical, treatment, and outcomes variables are summarized in Table 1. The cohort was predominantly male (88%) and white (91%). Median age-at-diagnosis was 57.8 years. Smoking status was never (43%), former (37%) and current (20%). Tumor subsite was primarily base-of-tongue (51%) and tonsil (41%). Median RT dose and total RT fractions were 70 Gray (Gy) and 33 fractions, respectively. Post-therapy feeding tube dependence was found in 99 patients (17%), and aspiration was reported in 96 (16%) patients at 6 months post-therapy. One quarter of patients (n = 152) reported RAD.

**Table 1.** Patient Characteristics. Legend: IQ: Inter-quantile, NOS: not otherwise specified, GPS: glossopharyngeal sulcus, RT: radiation therapy, IC: induction chemotherapy, CC: concurrent chemo-radiation, Gy: gray. Intra-cluster breakdowns of pathological characteristics are available in Appendix B.

| Characteristic | N (%) |
| --- | --- |
| **Gender** |  |
| Male | 512 (88) |
| Female | 70 (12) |
| **Median age at diagnosis (IQ Range)** | **57.8 (52.1-65)** |
| **Race** |  |
| White/Caucasian | 530 (91.1) |
| Black/African American | 17 (2.92) |
| Hispanic/Latino | 20 (2.92) |
| Asian | 7 (1.2) |
| Native American | 1 (0.66) |
| NOS | 7 (1.2) |
| **Smoking status** |  |
| Current | 118 (20) |
| Former | 216 (37) |
| Never | 248 (43) |
| **HPV Status** |  |
| Negative | 45 (8) |
| Positive | 360 (62) |
| Unknown | 177 (30) |
| **Tumor Sub-site** |  |
| Tonsil | 237 (41) |
| BOT | 295 (51) |
| Soft palate | 6 (1) |
| GPS | 11 (2) |
| NOS | 33 (6) |
| **Therapeutic combination** |  |
| Radiation alone (RT alone) | 74 (13) |
| Concurrent chemo-radiation (CC) | 308 (53) |
| Induction chemotherapy (IC) + RT alone | 53 (9) |
| IC+CC | 147 (25) |
| **Total RT dose, median (IQ Range)** | **70 (66-70) Gy** |
| **Total RT fractions, median (IQ Range)** | **33 (30-33)** |
| **Feeding tube at 6 months** |  |
| Y | 99 (17) |
| N | 483 (83) |
| **Aspiration rate at 6 months** |  |
| Y | 96 (16) |
| N | 486 (84) |
| **N Category (7th Edition)** |  |
| N1 | 71 (12) |
| N2a | 49 (8) |
| N2b | 311 (53) |
| N2c | 133 (23) |
| N3 | 18 (3) |
| **T-Category** |  |
| T1 | 131 (23) |
| T2 | 245 (42) |
| T3 | 121 (21) |
| T4 | 85 (15) |

In terms of LN spread and ignoring laterality, 63 distinct patterns of affected LNs were present in the cohort, with 6 patterns comprising 78% of the cases, as shown in Table 2. Of the patterns with greater than 10 cases, patients with bilaterally affected 2A-2B-3 levels showed the highest incidence of any toxicity (53%), while the group with unilaterally affected 2A-2B-3 regions had the lowest incidence (18%). Regions 2A and 2B were the most common affected sites, with 76% of patients showing unilateral spread and 19% showing bilateral spread in both nodes, whereas region 6 was not affected in any of the patients in the cohort.

**Table 2.** Affected lymph-node (LN) patterns and toxicity outcomes. Percentages of FT and AS are given as a percentage of the total patients with a given pattern. Legend: FT: Feeding tube. AS: Aspiration. 2A, 2B, 3, 4: lymph node level 2a, ab, 3 and 4.

| Pattern | Patient Count | FT | AS |
| --- | --- | --- | --- |
| Unilateral 2A-2B | 227 | 24 (11%) | 26 (11%) |
| Unilateral 2A-2B-3 | 125 | 16 (13%) | 13 (10%) |
| Bilateral 2A-2B | 37 | 10 (27%) | 4 (11%) |
| Unilateral 2A-2B-3-4 | 28 | 9 (32%) | 7 (25%) |
| Bilateral 2A-2B, Unilateral 3 | 19 | 6 (32%) | 4 (21%) |
| Bilateral 2A-2B-3 | 17 | 5 (29%) | 7 (41%) |
| Top 6 patterns total | 453 | 70 (15%) | 61 (13%) |
| Other (57) patterns | 129 | 29 (22%) | 25 (27%) |
| All 63 patterns | 582 | 99 (17%) | 96 (16%) |

The AUC for predicting feeding tube dependency, aspiration, and RAD using cluster labels for 2-5 clusters are shown in Table 3. Using 4 clusters yielded the best AUC performance for aspiration and overall RAD, defined here as presence of either feeding tube dependency or aspiration. Further merging the clusters into 3 groups yielded better AUC performance for predicting feeding tube toxicity, due to the fact that the 2 clusters that merged had different rates of aspiration but the same rate of feeding tube use. Using 5 clusters did not improve the AUC results. Incorporating N staging as an additional covariate in the regression model improved AUC performance in predicting aspiration, where N-stage and 3-4 clusters were the best performing model. However, for predicting feeding tube, using the 3 spatial clusters alone outperformed models that included N-stage. In the subsequent analysis, we used 4 clusters, as they performed best for overall dysphagia (RAD) in terms of AUC, and facilitated direct comparison against the current AJCC T and N staging systems, which describe, using 4 categories, the size of the tumor and spread to nearby tissue and lymph nodes.

**Table 3.** AUC scores (10-fold cross validation) for toxicity prediction using logistic regression from different numbers of lymph node clusters, with and without addition of N-staging. Models with the highest AUC for a given outcome are in bold. AUC performance is highest for 3 clusters and for 4 clusters, implying that those provide the best ranking of patients based on risk prediction.

|  | Cross-Validation AUC |  |  |  |  |  |
| --- | --- | --- | --- | --- | --- | --- |
|  | Feeding Tube |  | Aspiration |  | RAD |  |
| # Clusters | Our method | + N Category | Our method | + N Category | Our method | + N Category |
| Baseline: 1 cluster | 0.5 | 0.555 | 0.5 | 0.659 | 0.5 | 0.64 |
| 2 clusters | 0.577 | 0.572 | 0.591 | 0.675 | 0.591 | 0.646 |
| 3 clusters | **0.612** | **0.592** | 0.603 | 0.675 | 0.603 | 0.655 |
| 4 clusters | 0.595 | 0.567 | **0.621** | **0.677** | **0.621** | **0.656** |
| 5 clusters | 0.597 | 0.597 | 0.62 | 0.669 | 0.62 | 0.652 |

The 4 groups obtained from our clustering were significantly (p <.0001) with both feeding tube and aspiration rate at 6 months post-therapy (Appendix B). Clusters were also strongly correlated (p <.0001) with several other early risk factors: N-category, AJCC staging, T-category, tumor laterality, and total dose. Clusters were significantly correlated with HPV status (p <.01). No correlation was found between clusters and age (p >.05). Further, no correlation was found with tumor subsite (p >.05), possibly due to the prevalence of base-of-tongue cases. Breakdowns of demographics and toxicity by cluster are detailed in (Appendix B Table B1).

The 4 clusters leading to the highest AUC consist of 1 large, low risk cluster (Cluster 1), and 3 smaller, higher risk clusters that are most notably described by the spread and presence of bilaterally affected regions. To better explain the patient stratification to clinicians, we created a visual representation of the disease spread patterns in the clusters (Figure 2) [25]. The figure shows heat maps for the 4 clusters with the percentage of patients in that cluster who have specific LN levels affected, along with the incidence of different toxicity and high-risk clinical staging [26].

**Figure 2:**
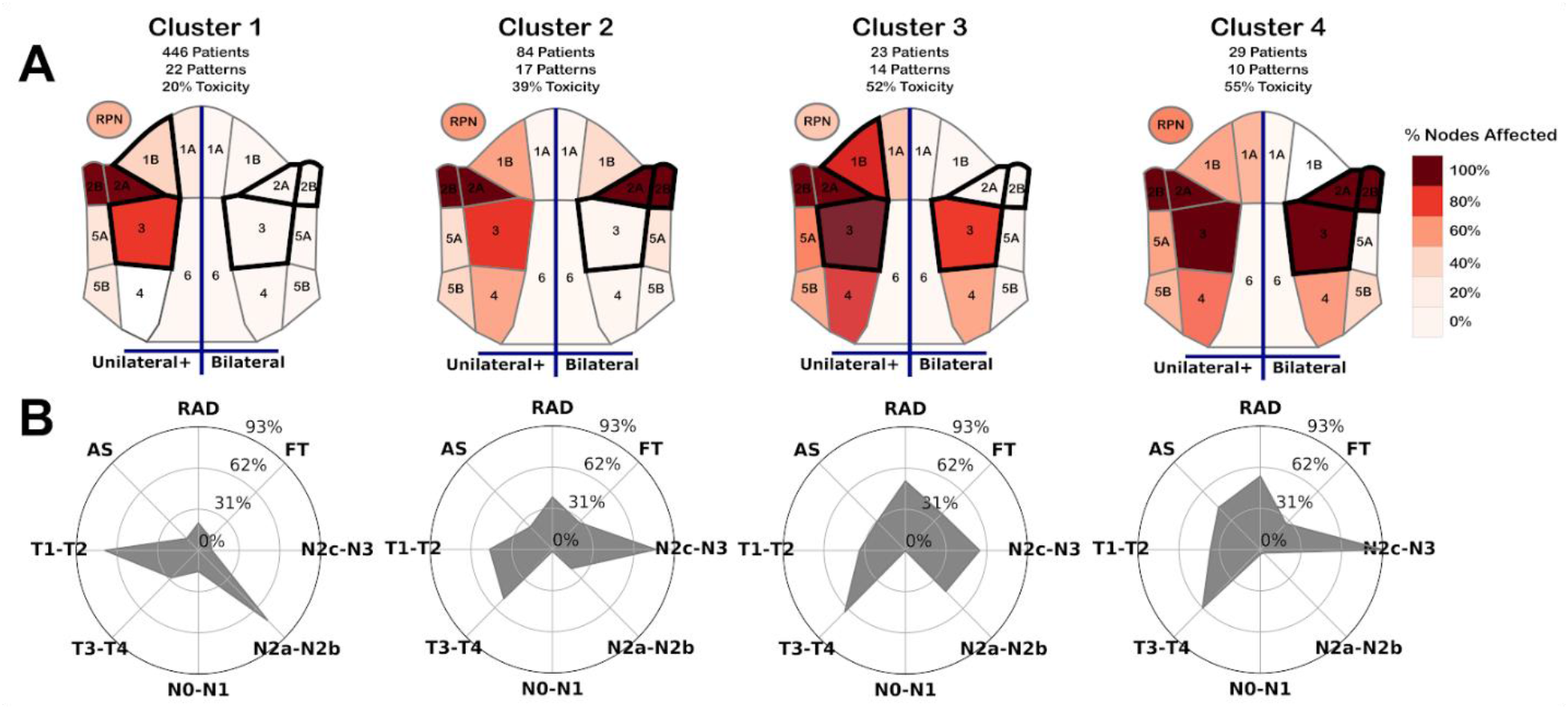
Visual representation of each cluster. (A) Heat map of nodal spread within each cluster. Left half of each map indicates % of patients in the cluster with at least one affected node on a specific level, while the right half encodes the percentage of patients with bilateral spread within a specific level. Regions outlined in black denote regions that are most discriminative for cluster membership, and could be used to determine if 99% of patients are within a given cluster. (B) Radar chart showing the % of patients in each cluster with a given toxicity or inclusion in a specific clinical staging category. FT: Feeding Tube, AS: Aspiration, RAD: Radiation-associated dysphagia, T1-2: T-category 1 or 2, T3-T4: T-category 3 or 4, N0-N1: N-category 0 or 1, N2a-N2b: N-category 2a or 2b, N2c-N3: N-category 2c or 3.

When using recursive partition analysis on the cohort, we found that baseline spread to levels 1B and 3, and bilaterality in levels 2A-2B-3 is sufficient to discriminate cluster spread for 99% of patients. Cluster 1 was the largest cluster, and included 446 patients and 22 patterns, including the 2 most common patterns: unilateral 2A-2B and unilateral 2A-2B-3, which cover 353 patients. This cluster is the lowest risk with 56 (12.5%) of patients with aspiration, 58 (13%) of patients with feeding tube insertion, and 91 (20%) of patients with overall RAD, which is lower than the overall cohort (26% RAD). This cluster had almost no bilaterally affected patterns, with the exception of a single patient, and had the lowest spread of any cluster.

Cluster 2 represents a moderate risk group, and includes 84 patients, 17 patterns, and 33 (39%) of patients had RAD. This cluster is characterized by having an above-average number of bilateral nodes affected; each pattern includes bilaterally affected levels 2A-2B. Cluster 2 has higher nodal spreads than cluster 1, but lower spreads than the other, higher risk clusters.

Cluster 3 is the smallest cluster, with 23 patients comprising 14 patterns. This cluster is best characterized as a high-risk cluster with 12 (52%) of patients showing RAD. Cluster 3 also has the highest risk of feeding-tube toxicity with 9 (39%) of patients. A minority of patients have bilaterally affected nodes in levels 3 or 4, but no other levels were bilaterally affected in this cluster, suggesting that this cluster largely represents the patterns with higher ipsilateral spread but few bilaterally affected regions.

Cluster 4 is composed of 29 patients and 10 patterns. 13 (44%) of patients had aspiration, 8 (27%) of patients required feeding tube, and 16 (55%) patients had RAD, making this the highest-risk cluster of aspiration and overall toxicity. All patterns were bilaterally affected in levels 2A-2B, and all but 1 pattern was bilaterally affected in level 3. Cluster 4 also has the highest spread on both the ipsilateral and contralateral sides, which likely contributes to its high toxicity risk.

Lower T and N stages were more prevalent in the large, low-risk group, with the lowest-risk Cluster 1 having the largest portion of patients in T-category I/II (71%), and AJCC category I/II (59%), while the highest-risk group Cluster 4 had the highest percentage of T-category 4 (38%) and AJCC summary stage IV (33%) among all 4 clusters. Cluster 1 (low risk) likewise had the largest portion of patients in N-categories 1 (15%), 2a (11%), and 2b (63%), the most patients with a total dose below 70 Gy (52%), and the most HPV positive patients (91%). A plurality of patients in clusters 2, 3, and 4 were in N-category 2c-3. Cluster 3 was more evenly divided between N-category 2b (43%) and N-category 2c (48%), whereas clusters 2 and 4 had larger portions of N-category 2c patients with 75% and 90% of patients, respectively, consistent with the fact that these clusters both have bilaterally affected nodes 2a-2b, and N-category 2c is composed of patients with bilaterally affected nodes.

After adjusting for age, smoking status, use of chemotherapy, HPV status, T category, and N category, LN clusters were significantly associated with RAD using a likelihood ratio test (p < 0.001). The addition of the LN clusters shows significant improvement when included in the model, even when T and N category are part of the model (Table 4).

**Table 4.** Akaike information criteria (AIC) and p-values from Likelihood ratio tests (LRT) for cluster labels in logistic regression models for predicting feeding tube, aspiration, and RAD. Lower AIC indicates better fit models after accounting for the number of covariates. LRT p-values <.05 indicates 95% confidence that cluster labels are meaningfully correlated with toxicity after accounting for other covariates. Legend: Age: Age >= 65; Smoke: Smoking Status (Never or Former/Current); Chemo: (Concurrent Chemo-radiation or Radiation Alone); HPV: HPV status (Positive, Negative, or Unknown); T: T-category (1-2 or 3-4); N: N-category (0-1 or 2-3).

|  | Feeding Tube |  |  | Aspiration Rate |  |  | RAD |  |  |
| --- | --- | --- | --- | --- | --- | --- | --- | --- | --- |
|  | Base model | Base model + LN Cluster |  | Base model | Base model + LN Cluster |  | Base model | Base model + LN Cluster |  |
| Model Covariates | AIC | AIC | LRT | AIC | AIC | LRT | AIC | AIC | LRT |
| Clinical (Age, Smoke, Chemo, HPV) | 519.1 | 508.6 | < .001 | 503.6 | 493.1 | < .001 | 645.1 | 627.1 | < .001 |
| Clinical + T-stage | 488.5 | 485.0 | < .05 | 477.2 | 472.3 | < .05 | 602.8 | 594.4 | < .01 |
| Clinical + T-stage + N-stage | 490.5 | **486.8** | < .05 | 475.2 | **467.5** | < .01 | 602.2 | **590.9** | < .001 |

## Discussion

Head and neck cancers account for nearly 3% of all malignancies in the U.S. with approximately 62,000 HNC cases diagnosed per year [2]. More than two-thirds of those diagnosed with HNC will survive 5 years or more if treated with locoregional curative therapy. However, almost all radiotherapy survivors will suffer from at least mild-to-moderate symptoms from head and neck radiation [14,27]. Recent phase III studies [28,29] suggest that concurrent chemoradiation will remain the standard for most locoregional head and neck cancers, and treatment protocols that minimize toxicity risk will remain essential. Thus, effective methods to predict radiation effects before radiation planning radiation, actively reducing the side effects to adjacent organs-at-risk (OARs) deliverable with current radiotherapy treatment devices represents a clinically key improvement in cancer care.

The vast majority of models utilized for radiation toxicity prediction consider individual organs-at-risk (OARs) independently, but fail to account for the anatomic proximity of distinct patterns of lymphatic spread, or to incorporate the fact that OAR dose is a function of proximity to tumor volumes [30]. As OPC typically exhibit substantial lymphatic involvement, pre-therapy predictive models for toxicity that are available before radiation planning are exceptionally valuable, as they can allow actionable therapy modification in terms of dose-modification and/or surgical neck management. Our ongoing aim is to determine whether a pre-radiotherapy planning spatial model could be incorporated into patient-education and shared-decision processes, based on an individual patient’s similarity to previously treated cases.

Concretely, this work introduces an unsupervised clustering technique to group patients based on the spread of diseased lymph nodes in OPC patients. We show that these clusters are significantly correlated with late radiation-associated dysphagia (RAD) after correcting for existing diagnostic information. Since our model relies on stratification using unsupervised clustering, it can allow for granular risk predictions for small populations of patients with uncommon or unique lymph node involvement. Thus, even before time intensive radiation plan optimization, an indication of the expected RAD can be estimated. Conclusively, this work shows that lymph node spread, determined at the time of diagnoses, can supply a clinically usable RAD risk assessment and guide toxicity-informed treatment decision making.

Most normal tissue complication probability (NTCP) models in practice rely on single dose-volume histograms for well-delineated organs-at-risks, or use dimensionality reduction to reduce complex dose distributions to a generalized uniform dose in order to predict late toxicity. In contrast, our model does not rely on the delineation of organs-at-risk or radiation dose optimization, which are time expensive, but instead uses the lymph-node spread as a precursor for late toxicity. Existing N-staging considers laterality, nodal gross tumor volume, and extranodal disease spread, yet does not consider any more complex anatomical information about the path of nodal spread, allowing discrimination based on chains of affected nodes. While our clusters are largely correlated with N-staging, we identified sub-patterns within these groups that are not captured by current staging considerations. The majority of the difference in the cohort presented as either in N-category 2b or lower (low risk) vs N-category 2c or higher (high-risk). However, our 3 high risk clusters were able to identify groups of higher risk patients within and across N-category 2b and 2c that allowed for more granular risk prediction not captured by N-category alone. Furthermore, after adjusting for age, smoking status, use of chemotherapy, HPV status, T-category, and N-category, the LN clusters were still significantly associated with RAD. Our technique uniquely considers 3-d anatomy of the lymphatic system in the oropharynx, allowing us to capture more granular spatial information beyond what is captured by current staging systems.

Interestingly, we found a disparity in risk within different clusters when comparing aspiration rate and feeding tube. Cluster 4 showed the highest percentage of RAD and a very high percentage of patients with aspiration. This is consistent with the group’s higher degree of bilaterally affected nodes and larger nodal spread than other nodes, as well as the higher T-category and AJCC stage, as patients with more severe disease are more likely to have adverse effects. However, feeding tube toxicity was most prevalent in cluster 3, suggesting that some underlying causes of feeding tube are not captured by overall disease spread or traditional AJCC staging. Notably, cluster 3 had no bilateral primary tumor or bilateral nodes in level 2A-2B, and only bilaterally affected nodes in levels 3-4. On the other hand, cluster 3 had the highest portion of tumors around the Tonsil (52%), while the other high-risk clusters had more tumors around the base of tongue (BOT). When ignoring bilaterality, cluster 3 also had the highest percentage of patterns with affected nodes in levels 1B, 4, 5A and 5B, indicating that cluster 3 may be separating a small set of patients with disease more localized to the lower regions of the neck, rather than the majority of patients with disease around levels 2A-2B. It may be the case then, that the presence of diseased lymph nodes in regions 4-5A-5B is indicative of feeding-tube toxicity risk separate from general disease spread or tumor size.

The resulting stratification allows for sophisticated, patient-specific combined representation of complex lymphatic disease spread in the head and neck. These results suggest that bilaterality, as well as spread to level 3 or 4, is correlated with clusters at high-risk of RAD, and is thus an indicator of patient risk of toxicity. However, the fact that our clusters, while heavily correlated with RAD, are largely subsumed in N-category 2b/2c, suggests that existing classifications methods fail to capture relevant spatial information in chains of affected lymph nodes. Our results also show that the maximum lymph node spread is a valuable covariate for stratifying patients, which is consistent with previous findings that suggest lymph node spread is a good measure of distance metastasis free survival~\cite{wu2019integrating}. Finally, our results show some distinction in patterns that are more correlated with aspiration rate vs feeding tube toxicity. While existing measures for survival are strongly related to measures for aspiration risk, such as bilaterality, lymph-node spread, and T-category, the specific location of tumors and affected lymph node chains lower in the neck may be underexplored indicators of late feeding tube insertion.

While this retrospective study considers a cohort with a large number of HNC patients and lymph node spread, it is not without limitations. First, our cohort consists primarily of white, male patients, and thus these results may not be reflected in more diverse demographics. Additionally, we consider only patterns present in our cohort. While this assumption is reasonable for a majority of patients, there may be different factors that may indicate RAD risk in patients with uncommon patterns of nodal spread that are not present in our cohort (e.g.,patients with nodal spread in level 6). Our results suggest that there is a small set of rare patterns with limited bilateral spread with higher risk of feeding tube toxicity. It is thus possible that there are spread patterns with bilaterally affected nodes in levels 3-5 that may be very high indicators of feeding tube insertion, but these patterns are rare, making it difficult to draw conclusions.

This work represents the first demonstration of a toxicity model incorporating spatial, inter-patient similarity metrics to predict radiation-associated dysphagia at 6-months postradiation in oropharyngeal cancer survivors. Future efforts will seek to integrate spatial features into visualization software for dynamic decision tools for multidisciplinary head and neck risk stratification, therapy selection, and patient education [6].

## Conclusion

In conclusion, this study demonstrates that clustering based on lymph nodes spread is associated with radiation-associated dysphagia, both aspiration toxicity and feeding tube dependency. Our anatomical representation of lymph node spread showed superior association to RAD compared to the current N-category classification. Our method relies only on discrete information on nodal spread at time of diagnosis, and thus does not require complex dose-planning or organ segmentation to determine RAD risk.

## Data Availability

The data are not available at this time.

## Appendix A: Multidimensional vector encoding for a patient's chain of affected lymph nodes

Each patient was represented using 3 different types of values:

1. Affected Nodes (10 values): Each of the 10 regions noted in the head were encoded as a unique covariate. Patients with disease on one side of the head were encoded with a 1, and patients with disease on both sides of the head in the given regions were encoded with a 2, while unaffected patients were encoded with a 0.
2. Node pairs (13 values): Where two regions were anatomically adjacent a ‘bigram’ covariate was created. Patients affected on both paired regions were encoded with either a 0, 1, or 2 depending on if the pair was unaffected, affected on one side of the head, or affected on both sides of the head, respectively.
3. Node spread (2 values): For each side of the head, a separate value encoded the minimum number of steps to move between the two furthest affected nodes in the head, equivalent to the ‘diameter’ of a graph of affected nodes. Since the RPLN is treated as a disconnected node, the maximum spread was increased by 1 when the RPLN was affected on the given side. Spread was treated as separate covariates for the side of the head with the primary tumor (ipsilateral) and the side opposite the primary tumor (contralateral). When the primary tumor (GTVp) crossed the midline of the head (bilateral), the side with the larger spread was treated as the contralateral side.

In total, we identified 10 regions in the head with 13 sets of adjacent regions, resulting in 25 covariates used to represent the affected LN levels of each patient. Figure 1 in the paper illustrates this encoding process for an example patient with bilaterally affected levels IIA-IIB and unilaterally affected level 3.

## Appendix B: Within-cluster distribution of patient covariates

Correlation between cluster cluster label and other relevant covariates. Highlighted cells indicate the cluster with the highest percentage of a given category. Percentages are given as relative to the known values in the relevant cluster, with the exception of the total patient counts, which are relative to the entire cohort.

**Table B1.** Distribution of clinical covariates and demographic data within each lymph node cluster.

|  | Cluster 1 |  | Cluster 2 |  | Cluster 3 |  | Cluster 4 |  | All |  | P-value (<) |
| --- | --- | --- | --- | --- | --- | --- | --- | --- | --- | --- | --- |
| Count | 446 | 77% | 84 | 14% | 23 | 4% | 29 | 5% | 582 |  |  |
| RAD | 91 | 20% | 33 | 39% | 12 | 52% | 16 | 55% | 152 | 26% | 0.00001 |
| Aspiration Rate | 56 | 13% | 20 | 24% | 7 | 30% | 13 | 45% | 96 | 16% | 0.00001 |
| Feeding Tube | 58 | 13% | 24 | 29% | 9 | 39% | 8 | 28% | 99 | 17% | 0.00005 |
| N Stage (7th Edition) |  |  |  |  |  |  |  |  |  |  |  |
| N0 | 1 | 0% | 1 | 1% | 0 | 0% | 0 | 0% | 2 | 0% | 0.00001 |
| N1 | 69 | 15% | 0 | 0% | 0 | 0% | 1 | 3% | 70 | 12% |  |
| N2a | 49 | 11% | 0 | 0% | 0 | 0% | 0 | 0% | 49 | 8% |  |
| N2b | 282 | 63% | 17 | 20% | 10 | 43% | 1 | 3% | 310 | 53% |  |
| N2c | 33 | 7% | 63 | 75% | 11 | 48% | 26 | 90% | 133 | 23% |  |
| N3 | 12 | 3% | 3 | 4% | 2 | 9% | 1 | 3% | 18 | 3% |  |
| T Stage |  |  |  |  |  |  |  |  |  |  |  |
| T1 | 122 | 27% | 5 | 6% | 2 | 9% | 2 | 7% | 131 | 23% |  |
| T2 | 195 | 44% | 35 | 42% | 6 | 26% | 9 | 31% | 245 | 42% |  |
| T3 | 83 | 19% | 23 | 27% | 8 | 35% | 7 | 24% | 121 | 21% |  |
| T4 | 46 | 10% | 21 | 25% | 7 | 30% | 11 | 38% | 85 | 15% | 0.00001 |
| Laterality (Primary Gross Tumor Volume) |  |  |  |  |  |  |  |  |  |  |  |
| Bilateral | 6 | 1% | 4 | 5% | 0 | 0% | 6 | 21% | 16 | 3% | 0.00001 |
| AJCC Category (8th Edition) |  |  |  |  |  |  |  |  |  |  |  |
| I | 44 | 14% | 0 | 0% | 0 | 0% | 0 | 0% | 44 | 11% |  |
| II | 217 | 69% | 40 | 69% | 9 | 53% | 6 | 40% | 272 | 67% |  |
| III | 30 | 10% | 10 | 17% | 3 | 18% | 4 | 27% | 47 | 12% |  |
| IV | 23 | 7% | 8 | 14% | 5 | 29% | 5 | 33% | 41 | 10% | 0.00002 |
| N/A | 132 |  | 26 |  | 6 |  | 14 |  | 178 |  |  |
| Total Dose to Primary Gross Tumor Volume |  |  |  |  |  |  |  |  |  |  |  |
| > 69 Gy | 231 | 52% | 63 | 75% | 18 | 78% | 20 | 69% | 373 | 64% | 0.00007 |
| HPV P16 Status |  |  |  |  |  |  |  |  |  |  |  |
| Negative | 27 | 9% | 8 | 14% | 5 | 29% | 5 | 33% | 45 | 11% | 0.006 |
| Positive | 288 | 91% | 50 | 86% | 12 | 71% | 10 | 67% | 360 | 89% |  |
| Unknown | 131 |  | 26 |  | 6 |  | 14 |  | 177 |  |  |
| Tumor subsite |  |  |  |  |  |  |  |  |  |  |  |
| BOT | 214 | 48% | 54 | 64% | 10 | 43% | 17 | 59% | 295 | 51% | 0.25 |
| GPS | 8 | 2% | 1 | 1% | 1 | 4% | 1 | 3% | 11 | 2% |  |
| NOS | 26 | 6% | 5 | 6% | 0 | 0% | 2 | 7% | 33 | 6% |  |
| Soft palate | 4 | 1% | 2 | 2% | 0 | 0% | 0 | 0% | 6 | 1% |  |
| Tonsil | 194 | 43% | 22 | 26% | 12 | 52% | 9 | 31% | 237 | 41% |  |

